# Predicting one-year postoperative functional status in contrast-enhancing glioma

**DOI:** 10.64898/2026.04.28.26351937

**Authors:** Eva Koderman, Marike R. van Lingen, Floris B. Tijhuis, Alexandros Ferles, Vera C. Keil, Ivar J.H.G. Wamelink, Sébastien Dam, Prejaas K. Tewarie, Matthan W.A. Caan, Philip C. De Witt Hamer, Linda Douw

**Affiliations:** Amsterdam UMC, Vrije Universiteit Amsterdam, Anatomy and Neurosciences, Amsterdam, Netherlands; Cancer Center Amsterdam, Brain Tumor Center, Amsterdam, The Netherlands; Amsterdam UMC, Vrije Universiteit Amsterdam, Radiology, Amsterdam, Netherlands; CERVO Brain Research Center, University of Laval, Quebec, Canada; Amsterdam UMC, Amsterdam Medical Center, Biomedical Engineering and Physics Department, Amsterdam, Netherlands; Amsterdam UMC, Vrije Universiteit Amsterdam, Neurosurgery, Amsterdam, Netherlands

**Keywords:** Glioblastoma, MRI, Karnofsky performance status, Machine learning, Neurosurgery

## Abstract

**Background and Objectives:** Preoperative prediction of functional outcomes in contrast-enhancing glioma could support surgical decision-making and patient counseling, yet most existing models incorporate histopathological or postoperative variables unavailable before surgery. Our objectives were to develop a preoperative-only prediction model for one-year functional status and evaluate the added value of MRI-based tumor characteristics beyond clinical predictors.

**Methods:** We conducted a retrospective cohort study of consecutive adults (≥ 18 years old) undergoing first resection of supratentorial contrast-enhancing glioma (WHO grade ≥ 2, histopathologically confirmed postoperatively) at a single center, with one-year follow-up. The primary outcome was functional status classified as mortality (Karnofsky Performance Score (KPS) = 0), functional dependence (KPS 10–60), or functional independence (KPS ≥ 70). In addition to clinical variables (age, sex, preoperative KPS, preoperative seizures), a deep learning tool was used to extract structural MRI-based tumor characteristics as predictors. A machine-learning model was developed and conformal prediction was applied to stratify patients by prediction confidence level.

**Results:** 552 patients were included (median age: 60 years, range: 18–84; median contrast-enhancing volume: 24 mL, IQR: 10–43; median preoperative KPS: 80, range: 30–100; retrospectively confirmed 88% glioblastoma). Most MRI-based predictors did not improve performance as the best-performing model included three predictors: age at diagnosis, contrast-enhancing volume, preoperative KPS. Bootstrapped areas under the curves were 0.77 (95% confidence interval 0.70–0.84) for mortality, 0.64 (0.52–0.77) for functional dependence, and 0.71 (0.63–0.79) for functional independence. F1 scores per class were 0.65, 0.24, 0.65, respectively. Conformal prediction provided reliable predictions for 18% patients, moderate uncertainty for 57%, and identified 25% with genuinely unpredictable outcomes.

**Discussion:** Our preoperative machine-learning model predicted one-year functional status in contrast-enhancing glioma with functional independence being the most reliably classified outcome (ROC-AUC = 0.77, F1 score = 0.65) and functional dependence the most challenging to predict (ROC-AUC = 0.64, F1 score = 0.24). A small set of three preoperative predictors drove model performance, supporting generalizability to broader patient populations. Our open-source model enables individualized risk stratification and may help clinicians identify patients with uncertain prognoses warranting more intensive preoperative counseling or follow-up planning.

## Introduction

Glioma is typically identified using magnetic resonance imaging (MRI), which can show a contrast-enhancing mass, a potential radiological indicator for a glioblastoma.^1^ Resection is the primary treatment when feasible, followed by concurrent chemo-radiotherapy completed within nine months.^2^ Permanent loss of brain function and adjuvant treatment-related adverse effects can result in functional dependence.^3^ Assessing outcomes one year after resection provides a pragmatic clinical landmark.^4^

This can be done using the Karnofsky Performance Status (KPS)^5^, with a score of 70 representing important boundary between functional independence and dependence. In glioma, a score below 70 is associated with poorer prognosis^6^ and quality of life.^7^ Such outcomes are difficult to predict objectively and are often based on clinicians’ prior experience and hope, which tend to overestimate postoperative functional status.^8^

Machine learning (ML) offers a promising quantitative alternative for patient-specific outcome prediction. ML models, like eXtreme Gradient Boosting (XGBoost), can be directly interpreted, revealing which predictors drive specific predictions.^9^ Additionally, ML can be combined with an automatic approach to extract quantifiable tumor-related features from clinically obtained routine MRIs.^10^ This is done by a deep learning-based automatic tumor segmentation, which is used to generate an automatic report detailing MRI-based tumor characteristics, such as tumor location, multifocality, laterality, and others.^11^ While such tumor characteristics are readily available to clinicians, deep learning-based extraction can be used to scale their derivation across a large research cohort.

Long-term postoperative KPS prediction remains challenging as only information before surgery is available without the histopathological report. Current models focus on predicting outcomes up to six months to evaluate potential continuation with chemo-radiotherapy.^12–14^ However, most include intraoperative or postoperative predictors, limiting their clinical preoperative usability. Moreover, to assess the complete treatment effect, predictions should be done for minimum nine months postoperatively. This is crucial in contrast-enhancing glioma, where poor prognosis demands careful treatment planning to avoid overtreatment.^1,3^

Therefore, this study aimed to: (1) predict postoperative functional status in contrast-enhancing glioma at around one year after surgery, (2) relying exclusively on preoperative predictors, and (3) evaluate the added value of MRI-based tumor predictors alongside clinical data. Additionally, we explored the use of Mondrian conformal prediction (MCP), which can provide added insight into prediction uncertainty and patient stratification confidence levels as an extension to the model.^15–17^

## Methods

### Patients

This single-center retrospective cohort study included all eligible consecutive patients at the Amsterdam UMC, location VUmc, from the hospital glioma database (IMAGO). Patients were included if they 1) had a supratentorial infiltrative glioma with at least 1 voxel labeled as gadolinium contrast-enhancing tumor component, 2) were ≥18 years old, and 3) underwent their first tumor resection between 2010 and 2022. Patients undergoing biopsy rather than resection have a different clinical trajectory and were therefore not included.^18^ Approval was received from the institutional medical ethical committee of the VUmc (Vumc_2021-0437). Informed consent was waived.

Patient exclusion steps are summarized in Supplementary Figure 1. Patients were excluded if T1-weighted preoperative scans were unavailable or failed visual quality inspection, as required for further MRI processing. Finally, those without postoperative KPS at around one year, or without enough clinical information to estimate it with, were excluded.

### Outcome Measure

The main outcome in this study was KPS categorized into mortality (KPS = 0), functional dependence (KPS 10–60), and functional independence (KPS ≥ 70). For surviving patients, the KPS score was retrieved at postoperative one year within a range of three months. If not explicitly documented in the medical record or available databases, two trained clinical researchers retrospectively assigned the KPS value based on patient medical notes from the same time frame. To minimize interrater variability, part of the retrospectively assigned KPS scores rated by the two researchers were cross-checked, and discrepancies were resolved by consensus and scoring choices standardized.

### Predictor Variables

Predictors were chosen based on data availability from preoperative clinical routine visits. Clinical predictors included sex (categorical), history of preoperative seizures (categorical yes or no), age (continuous), and preoperative KPS score (ordinal). Similar to postoperative KPS, the preoperative KPS was retrospectively assigned in patients where this information was not available from the medical records.

MRI-based predictors were derived as follows. First, the automatic tumor segmentation was obtained using the PICTURE toolbox, which is a deep learning algorithm trained on more than 1200 manual glioma segmentations.^19^ All available MRI sequences were used (gadolinium contrast-enhanced T1-weighted, T1-weighted, T2-weighted, fluid-attenuated inversion recovery), with gadolinium-enhanced T1-weighted imaging required in all patients. The output was a tumor mask with three components — necrotic core, contrast-enhancing tumor, and T2 hyperintensity. The volumes of each of these components were calculated in the Montreal Neurological Institute standard space (MNI ICBM 2009b NLIN Asymmetric). This was done by counting the voxels within each component label, multiplying by the voxel volume, and converting to milliliters (mL). Each component was thus represented by a single volume value (mL).

The three tumor mask components were subsequently combined into a single tumor mask for further analysis with the GSI-RADS tool.^11^ Feature selection was performed based on expert consultation regarding variable relevance. The chosen features from the GSI-RADS output included the tumor laterality index (ranging from −1 for left-sided to +1 for right-sided), multifocality (categorical yes or no), and expected resection percentage (derived from probability maps^20^). Additionally, we included tumor volume overlaps with various brain structures, calculated as the number of tumor voxels within each anatomical structure expressed as a percentage of the total tumor volume. For clarity, the tumor overlap values of individual anatomical structures were summed within the following categories: cognitive networks (salience/ventral attention, dorsal attention, frontoparietal/control, default mode network according to the 7-network Yeo parcellation^21^), sensory limbic networks (visual, somato-motor, limbic network), frontal lobe, temporal lobe, parietal lobe, occipital lobe, and white matter tracts (described in Table 1). This aggregation approach was previously described.^22^

**Table 1:**
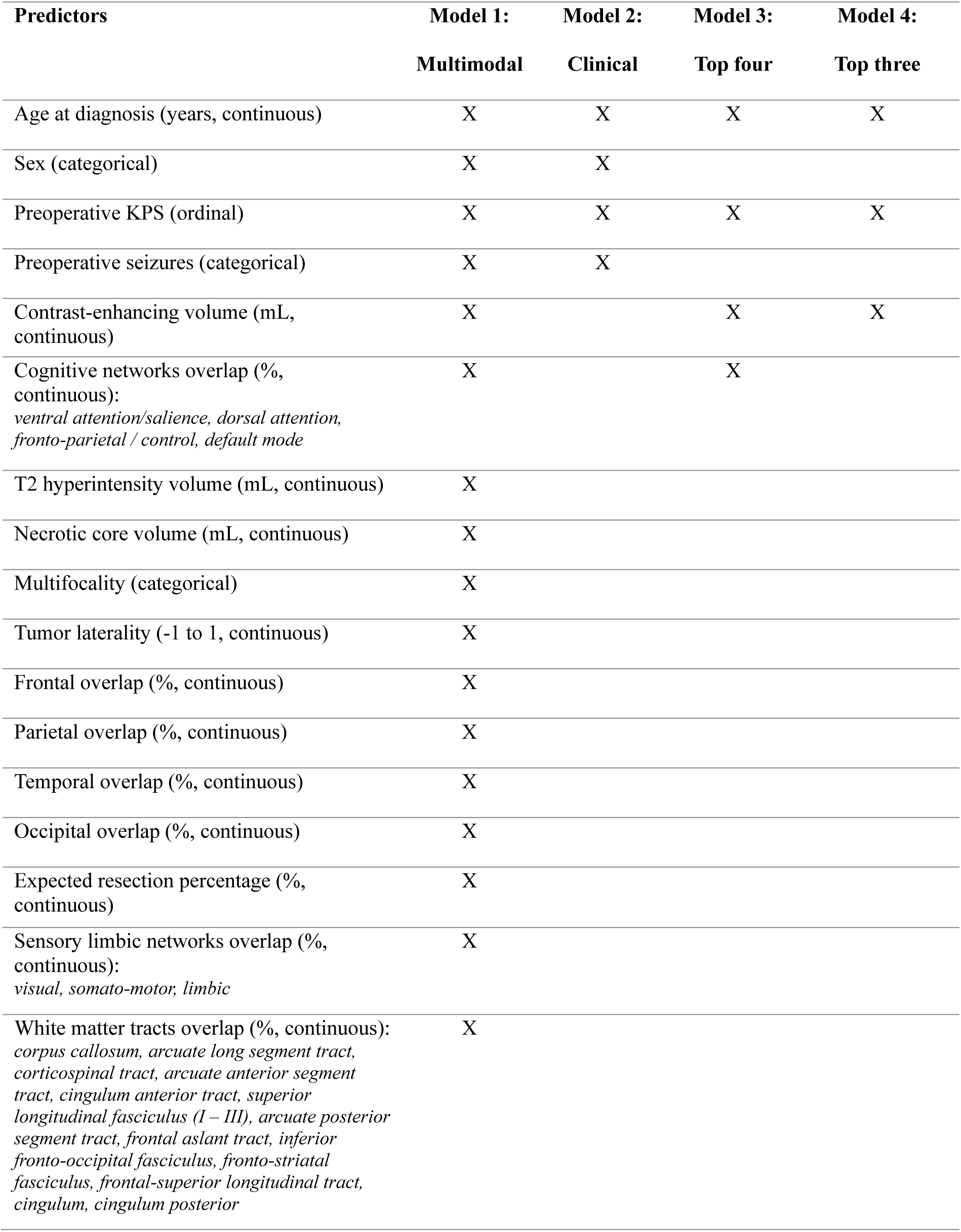
Different predictors and their corresponding aggregated structures used across the models.

| Predictors | Model 1:<br>Multimodal | Model 2:<br>Clinical | Model 3:<br>Top four | Model 4:<br>Top three |
| --- | --- | --- | --- | --- |
| Age at diagnosis (years, continuous) | X | X | X | X |
| Sex (categorical) | X | X |  |  |
| Preoperative KPS (ordinal) | X | X | X | X |
| Preoperative seizures (categorical) | X | X |  |  |
| Contrast-enhancing volume (mL, continuous) | X |  | X | X |
| Cognitive networks overlap (% continuous):<br>*ventral attention/salience, dorsal attention, fronto-parietal / control, default mode* | X |  | X |  |
| T2 hyperintensity volume (mL, continuous) | X |  |  |  |
| Necrotic core volume (mL, continuous) | X |  |  |  |
| Multifocality (categorical) | X |  |  |  |
| Tumor laterality (-1 to 1, continuous) | X |  |  |  |
| Frontal overlap (% continuous) | X |  |  |  |
| Parietal overlap (% continuous) | X |  |  |  |
| Temporal overlap (% continuous) | X |  |  |  |
| Occipital overlap (% continuous) | X |  |  |  |
| Expected resection percentage (% continuous) | X |  |  |  |
| Sensory limbic networks overlap (% continuous):<br>*visual, somato-motor, limbic* | X |  |  |  |
| White matter tracts overlap (% continuous):<br>*corpus callosum, arcuate long segment tract, corticospinal tract, arcuate anterior segment tract, cingulum anterior tract, superior longitudinal fasciculus (I – III), arcuate posterior segment tract, frontal aslant tract, inferior fronto-occipital fasciculus, fronto-striatal fasciculus, frontal-superior longitudinal tract, cingulum, cingulum posterior* | X |  |  |  |

### Machine-Learning Model

Model development and reporting was done in accordance with the TRIPOD guidelines.^23^ The analysis was prospectively preregistered (<u>10.17605/osf.io/f29xm</u>). This study addressed a three-class ordinal classification problem (mortality, functional dependence, functional independence) using an XGBoost-based ordinal regression model (Python 3.10, scikit-learn 1.2.2).^24^

### Model Development

The dataset was split into 70% development set and 30% validation set to maintain adequate sample size in both sets while ensuring sufficient representation of all outcome categories. Stratified sampling was performed based on a composite label combining event status (alive at last follow-up or deceased with timing: <12 months, 12-15 months, or ≥15 months) and postoperative KPS (KPS <70 or KPS ≥70). To ensure stable allocation, the presence of rare strata was assessed and none were identified.

Ordinality was imposed by two underlying binary XGBoost classifiers corresponding to adjacent outcome thresholds. The first classifier distinguished mortality from survival (Classifier 1: mortality versus functional dependence and independence), whereas the second distinguished functional independence from mortality or functional dependence (Classifier 2: functional independence versus functional dependence and mortality). During inference, the probability of mortality was calculated as 1 – probability_survival_ from Classifier 1, the probability of functional independence was taken directly from Classifier 2, and the probability of functional dependence was derived as probability_survival_ – probability_functional_independence_. The resulting probabilities were normalized by clipping negative values to zero and rescaling to sum to one. Given this model choice and predictors, no feature scaling or imputation of predictors were implemented.

The development procedure employed a nested cross-validation strategy, with a stratified 10-fold outer loop for performance estimation and a stratified 3-fold inner loop for hyperparameter tuning. Hyperparameter optimization was performed using Bayesian optimization (BayesSearchCV, scikit-optimize 0.9.0) to maximize the weighted one-vs-rest area under the receiver operating characteristic curve (ROC-AUC) score, which accounts for class imbalance. Class imbalance was further addressed by sample weights computed inversely proportional to class frequencies. Additionally, samples from the class with the lowest count (functional dependence) were scaled by a multiplicative factor tuned as a hyperparameter, with range chosen empirically to balance minority class detection against excessive misclassification of the majority classes. All weights were then normalized to unit mean. Details of the hyperparameter search space are provided in Supplementary Figure 2.

### Model Selection

Predictors used in models are summarized in Table 1. Initially, two models were trained: one using clinical predictors and a multimodal using clinical and MRI-based predictors. Shapley (SHAP) values were computed, which represent each feature’s contribution to the model’s log-odds output, with positive values increasing and negative values decreasing the outcome predicted probability.^25^ Mean absolute SHAP values across classifiers were also computed to assess global feature importance independent of direction. To evaluate whether a reduced predictor set could match or exceed the multimodal performance, four models, with identical underlying ordinal XGBoost architecture, were compared: multimodal, clinical, top four, and top three predictors.

The best-performing model among these four was selected as the final model and evaluated on the validation set. The decision for the final model was based on the following metrics across folds: F1-weighted score (0-1, higher is better), mean absolute error (MAE; 0-∞, lower is better), quadratic weighted kappa (QWK; −1 to 1, higher is better), Matthews correlation coefficient (MCC; −1 to 1, higher is better, with 0 indicating random prediction), and ROC-AUC (0-1, higher is better, with 0.5 indicating chance performance). Additionally, we considered optimizing clinical usability by favoring fewer input variables readily available from routine clinical assessments.

### Model Validation

The final model was evaluated on the validation set. Prior to final testing, the optimal hyperparameters for the model were determined from the development set. For each fold of the nested cross-validation, the best hyperparameters were recorded, and the final hyperparameter values were chosen as the median across all folds. These final parameters were used to train the model on the full development set before evaluating performance on the validation set. Sample weights were again calculated as the inverse of each class’s frequency to account for class imbalance, consistent with the approach used during model training. The trained model was applied to the validation set to generate predicted class probabilities.

Model performance on the validation set was assessed using multiple metrics. Discrimination was quantified using weighted one-vs-rest ROC-AUC, complemented by confusion matrices and additional classification metrics (F1 score, QWK, MCC). Metrics were bootstrapped (500 iterations) to derive metric-specific confidence intervals. An external validation cohort was not available for this study.

### Mondrian Conformal Prediction

To provide statistically valid uncertainty quantification, MCP was applied as model extension. Out-of-fold prediction probabilities from the development set were used to compute class-specific nonconformity scores, defined as 1 – predicted probability_true_outcome_. The empirical distribution of these scores within each class was used to determine class-specific threshold at the (1-α) quantile with finite-sample correction (α=0.15), ensuring that prediction sets would contain the true outcome in at least 85% of cases. For each new patient, an outcome was included in the prediction set if its nonconformity score fell below the corresponding class threshold. The calibrated thresholds were evaluated on the validation set to confirm that the target coverage was achieved.

The primary output of MCP is the prediction set, which contains one or more classes for each patient based on the calibrated thresholds. Prediction set size stratified patients by certainty: single-outcome sets indicated reliable predictions, two-outcome sets reflected moderate uncertainty, and three-outcome sets identified genuinely ambiguous cases where baseline predictors provide insufficient discrimination, warranting close clinical monitoring of these patients.

## Results

### Patient Characteristics and Outcome Measure

We included 552 eligible patients (N total = 650, N excluded = 98, Supplementary Figure 1). Patients had the following main characteristics (Table 1, Supplementary Table 1): age at diagnosis (median, range; 60 years, 18–84), contrast-enhancing volume (median, IQR; 24 mL, 10–43), preoperative KPS (median, range; 80, 30–100). Most patients had all types of scans available (N = 496, Supplementary Table 2). Glioma subtypes were confirmed through retrospective review of histopathology, revealing glioblastomas (N = 487; 88%), astrocytomas (N = 26; 5%), oligodendrogliomas (N = 35; 6%), and other gliomas (N = 4; <1%). Moreover, retrospective verification of treatment procedures revealed that most patients underwent both chemotherapy and radiotherapy (N = 424; 77%), with some patients undergoing chemotherapy only (N = 18; 3%), radiotherapy only (N = 43; 8%), or no treatment (N = 68; 12%) (Supplementary Table 3). Tumor frequency maps are provided in Supplementary Figure 3.

Postoperative KPS of 0 (mortality) was assigned in 221 (40%) patients, a KPS of 10–60 (functional dependence) in 68 (12%) patients, and a KPS of ≥ 70 (functional independence) in 263 (48%) patients. In some patients, assigning an exact KPS score was not possible due to limited detail in the patient medical records (preoperative KPS N = 5; postoperative KPS N = 13). However, the information was sufficient to determine whether KPS would be assigned to either the functionally dependent or functionally independent class. These patients were included in Table 2 with the maximum value of the range (a KPS of 60 or 100). This did not affect model predictions, as the model used KPS ranges rather than specific KPS values.

**Table 2:** Patient characteristics.

| **Characteristic** | **All**<br>**(N = 552)** | **Mortality**<br>**(N = 221; 40%)** | **Functional**<br>**dependence**<br>**(N = 68; 12%)** | **Functional**<br>**independence**<br>**(N = 263; 48%)** |
| --- | --- | --- | --- | --- |
| Age, median years<br>(min - max) | 60 (18 – 84) | 65 (32 – 84) | 61.5 (25 – 80) | 54 (18 – 82) |
| Preoperative KPS,<br>median (min - max) | 80 (30 – 100) | 80 (30 – 100) | 80 (30 – 100) | 90 (30 – 100) |
| Enhancing component<br>volume (ml), median<br>(Q1 – Q3) | 24<br>(10 - 43) | 33<br>(20 - 49) | 18<br>(7 - 35) | 16<br>(4 - 38) |
| Tumor overlap with<br>cognitive networks<br>(%), median (Q1 – Q3) | 36<br>(29 - 43) | 34<br>(27 - 40) | 35<br>(30 - 41) | 38<br>(32 - 47) |
| Tumor laterality,<br>left / right / bilateral (%) | 217 (39) /<br>299 (54) /<br>36 (7) | 79 (36) /<br>132 (60) /<br>10 (5) | 29 (43) /<br>36 (53) /<br>3 (4) | 109 (41) /<br>131 (50) /<br>23 (9) |
| Glioblastoma/<br>Astrocytoma/<br>Oligodendroglioma/<br>other glioma (%) | 487 (88) /<br>26 (5) /<br>35 (6) /<br>4 (<1) | 216 (98) /<br>1 (<1) /<br>4 (2) / | 66 (97) /<br>1 (1) /<br>1 (1) / | 205 (78) /<br>24 (9) /<br>30 (11) /<br>4 (2) |

Missingness was present in the following predictors: preoperative seizures (N = 5; 1%), preoperative KPS (N = 1; 0.2%), T2 hyperintensity volume (N = 6; 1%) and necrotic core volume (N = 14; 3%). Missing predictor values were not imputed, as XGBoost handles missingness natively.

### Machine-Learning Model

#### Model Selection

The dataset was divided into a development (N = 386, 70%) and a validation set (N = 166, 30%). Models were trained on the development set, and their performances were compared to find the best performing one (Table 1; list of predictors used across the models, Supplementary Table 4; comparison of performances across models, Supplementary Figure 4; F1 score per class per model).

Figure 1 shows the SHAP values for the global predictor importance across both classifiers, as well as the two binary classifiers, indicating the directionality of each predictor on the model output. Global SHAP analysis on the multimodal model identified four key predictors: age at diagnosis, contrast-enhancing volume, preoperative KPS, and tumor overlap with the cognitive networks, ranked by importance. These were further explored in separate models, which comprised of top four predictors and top three predictors (excluding tumor overlap with the cognitive networks) as described in Table 1.

**Figure 1:**
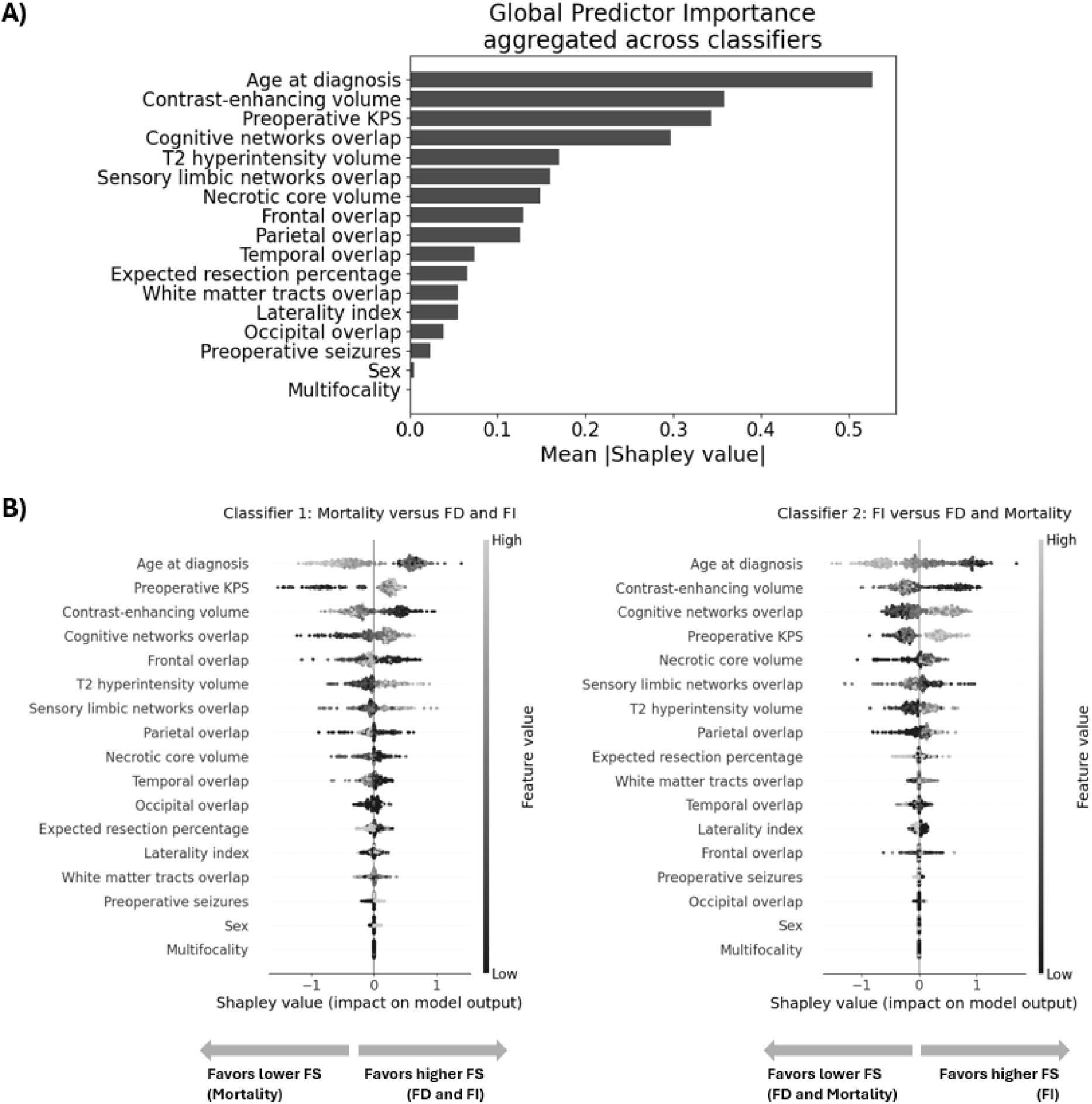
Shapley values derived from the multimodal model trained on the development set (N = 386). In both panels, features are ordered based on importance. A) The global Shapley values plot summarizes feature importance by displaying the mean absolute SHAP value of each feature across all samples and classifiers, quantifying each feature’s average contribution to model predictions regardless of direction. The order of features shows the order of importance. B) Shapley values for each of the two-underlying binary XGBoost classifiers correspond to adjacent outcome thresholds. The first classifier distinguished mortality from functional dependence and independence, whereas the second distinguished functional independence from mortality and functional dependence. Abbreviations: FD = Functional dependence, FI = Functional independence, FS = Functional status.

The multimodal model demonstrated the worst performance for the functional dependence class in the ROC-AUC (0.56) and F1 score (0.18). The models with top three and top four predictors showed similar performance, but the model with top four predictors indicated higher variability across folds in almost all evaluation metrics (predictors description in Table 1; results in Supplementary Table 4, Supplementary Figure 4). The top three predictors model (F1 weighted = 0.58, QWK = 0.42, MCC = 0.32) outperformed the clinical only model (F1 weighted = 0.58, QWK = 0.36, MCC = 0.29). Additionally, to increase clinical usability, lower number of predictors was preferred. Therefore, the final chosen model was the top three model, which was further evaluated on the validation set.

#### Model Evaluation

Figure 2 shows the final model’s discriminatory performance using the top three globally most important features evaluated on the validation set (age at diagnosis, contrast-enhancing volume, preoperative KPS). ROC–AUC showed the best discrimination for mortality (0.77), intermediate performance for functional independence (0.71), and poorest for functional dependence (0.64). The confusion matrix suggested an overlap in feature space, rather than a systematic bias toward a single wrong class.

**Figure 2:**
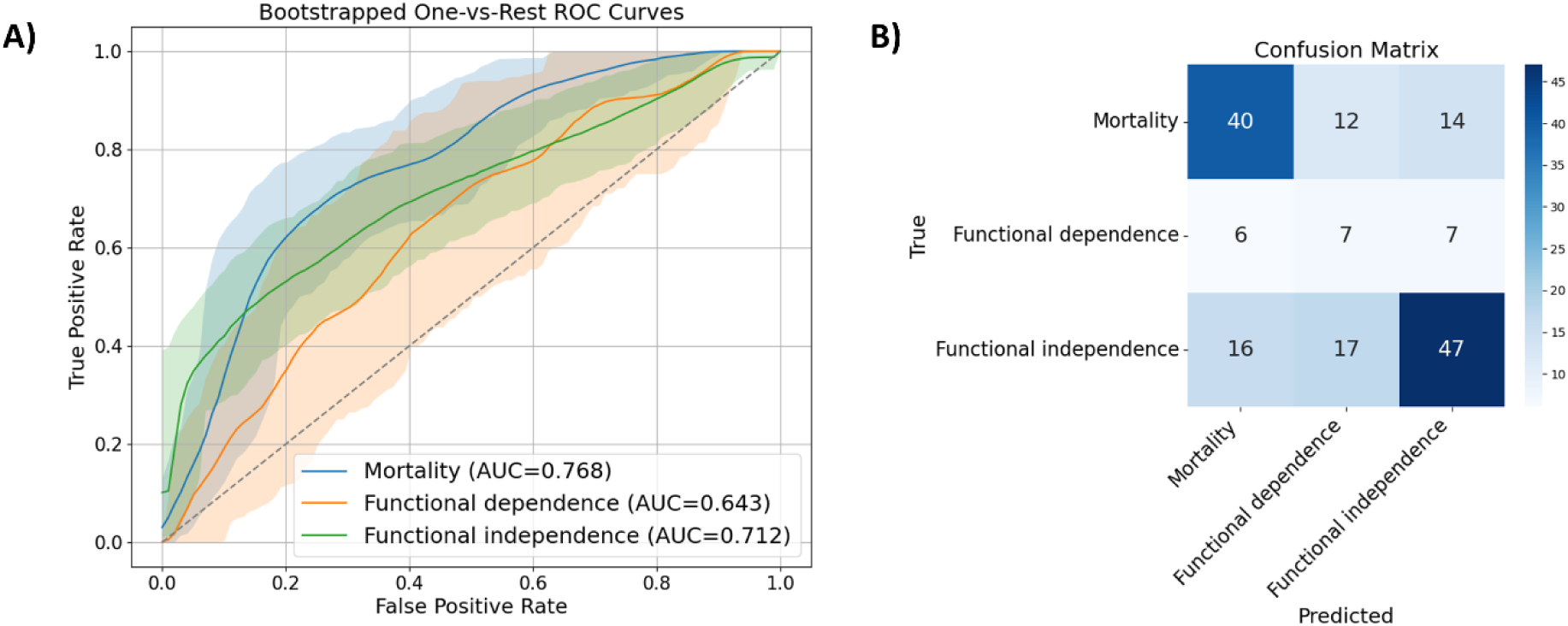
Discriminatory performance of the final model trained with age at diagnosis, contrast-enhancing volume, and preoperative KPS evaluated on the validation set (N = 166). A) ROC-AUC one-vs-rest curves show how well the model discriminates one class from the remaining classes - each class is treated as the positive class with the other two classes combined as the negative class. B) Confusion matrix shows true versus predicted class labels for the model. Abbreviations: ROC-AUC = Receiver Operating Characteristic - Area Under the Curve.

Table 3 shows the final model’s performance on the validation set. Mortality (F1 score = 0.62) and functional independence (F1 score = 0.64) demonstrated a fairly good balance between precision and recall with reasonably accurate classification. The functional dependence showed poor balance between precision and recall (F1 = 0.25), with many misclassifications. The global performance metrics across classes were QWK = 0.41, indicating moderate agreement with true ordinal classes, and MCC = 0.31, indicating modest overall predictive ability accounting for class imbalance.

**Table 3:** Model performance evaluation on the validation set. Abbreviations: ROC-AUC = Receiver operating curve – area under the curve; CI = confidence interval.

| **Metric** | **Validation set**<br>**(N = 166)** |
| --- | --- |
| **F1 score (95% CI)** |  |
| Mortality | 0.62 (0.52 – 0.72) |
| Functional dependence | 0.25 (0.11 – 0.40) |
| Functional independence | 0.64 (0.53 – 0.72) |
| **ROC-AUC (95% CI)** |  |
| Mortality | 0.77 (0.70 – 0.84) |
| Functional dependence | 0.64 (0.52 – 0.76) |
| Functional independence | 0.71 (0.63 – 0.79) |
| **Quadratic weighted kappa**<br>**(95% CI)** | 0.41 (0.27 – 0.54) |
| **Matthew’s correlation**<br>**coefficient (95% CI)** | 0.31 (0.20 – 0.43) |

Overall, the model achieved moderate accuracy and agreement, with a fairly good performance for mortality and functional independence and less than optimal performance for functional dependence. The discriminative ability (ROC-AUC) aligned with class-wise accuracy metrics (F1), demonstrating consistent model performance with difficulty in distinguishing the intermediate functional dependence category. Additionally, the model performance was comparable between the development and validation set, suggesting minimal overfitting and good generalizability within this patient population (Table 3, Supplementary Table 5).

#### Model Explainability

Figure 1 shows the global and classifier-specific predictors, ranked by contributions to predictions. The most significant predictor was age at diagnosis (mean |SHAP| = 0.53). It was followed by contrast-enhancing volume (mean |SHAP| = 0.36), preoperative KPS (mean |SHAP| = 0.34), and tumor overlap with cognitive networks (mean |SHAP| = 0.30). The remaining predictors had mean |SHAP| values below 0.3, with multifocality showing negligible contribution to model predictions (mean |SHAP| ≈ 0). The directionality of the top four predictors with SHAP values above 0.3 was further inspected per classifier.

Both binary classifiers (Classifier 1: mortality versus functional dependence and independence and Classifier 2: functional independence versus functional dependence and mortality) identified younger age, higher preoperative KPS, and smaller contrast-enhancing volume as the strongest predictors of survival and functional independence. The second classifier additionally identified that bigger tumor overlap with cognitive networks was associated with functional independence, ranking above the preoperative KPS. Additional analysis demonstrated a negative correlation (r = −0.54) between the tumor overlap with the cognitive and the sensory limbic networks (Supplementary Figure 5).

#### Mondrian Conformal Prediction

The implementation of MCP is visualized using three example patients in Figure 3, and the calibration and validation results are summarized in Supplementary Table 6. Class-specific thresholds for mortality, functional dependence, and independence were 0.81, 0.85, 0.80, meaning that the minimum probability per class to be included in the set was 19%, 15%, 20%, respectively. Overall, thresholds were similar across classes, although the functional dependence class had the highest thresholds, meaning it was the most difficult to predict confidently. The achieved coverage rates on the validation set for mortality, functional dependence, and functional independence were 91%, 85%, and 80%, respectively. This closely followed the target 85% coverage (α=0.15), with functional dependence showing slight under-coverage.

**Figure 3:**
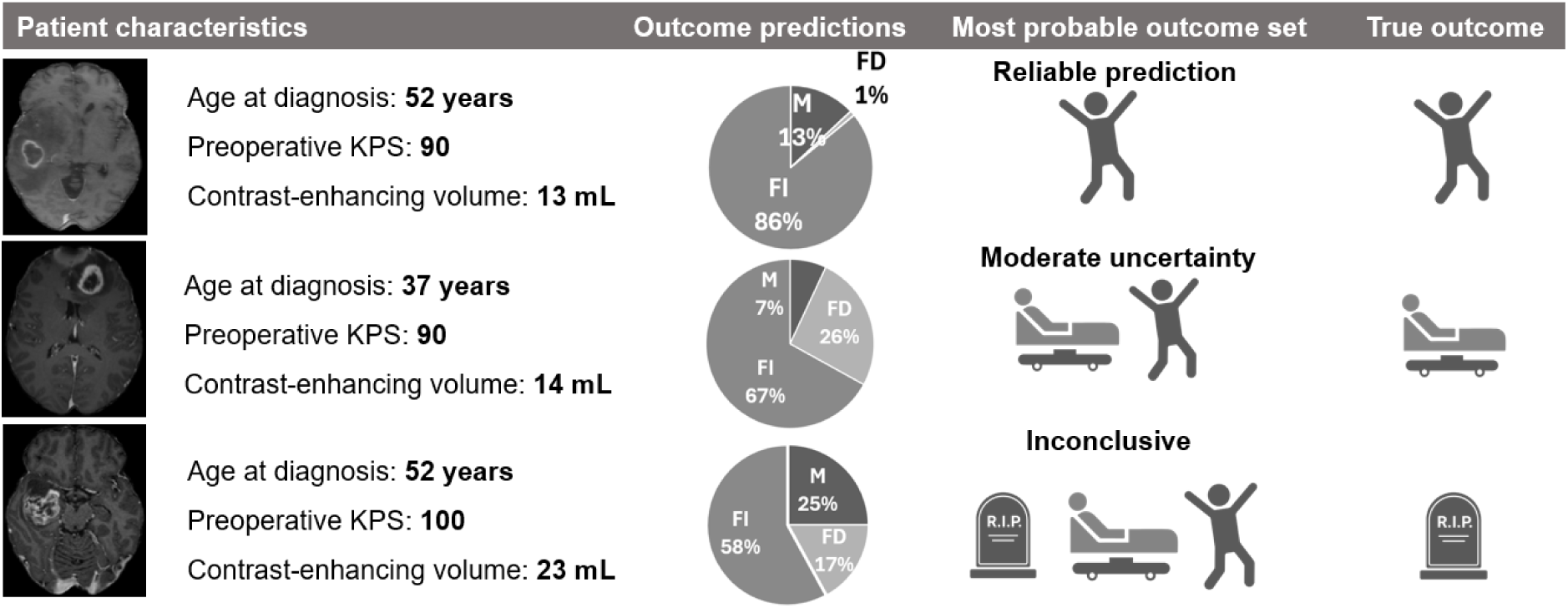
Example patients with outcome predictions using Mondrian conformal prediction (MCP). The model assigns point probabilities to three outcomes; MCP then generates an outcome set guaranteed to contain the true outcome in 85% of patients with similar characteristics. Set size reflects prognostic certainty with three-outcome sets clearly showing insufficient discriminability in which the model abstains from a confident prediction.

Of the validation set, 18% of patients had single-outcome sets, 57% had two-outcome sets, and 25% had three-outcome sets. Feature space analysis revealed that uncertain patients (three outcomes) showed predominantly middle-range predictor values (Supplementary Figure 6A). True outcome distributions were balanced across prediction set sizes (Supplementary Figure 6B), confirming that flagged uncertain cases reflect genuine unpredictability rather than model underperformance.

#### Model Deployment

To enhance clinical usability, the model was integrated into an open-source, user-friendly, web-based application available at https://glioma-kps-prediction.streamlit.app/.

## Discussion

Predicting postoperative functional outcome in glioma is inherently challenging, as molecular markers, histology, and postoperative treatment information are unavailable prior to resection. We developed a prediction model relying on preoperative predictors alone. The model predicts the one-year postoperative functional status consisting of mortality (ROC-AUC = 0.76), functional dependence (ROC-AUC = 0.64), and functional independence (ROC-AUC = 0.71) using age at diagnosis, preoperative KPS, and contrast-enhancing volume. Notably, additional tumor characteristics extracted from the MRIs resulted in worse functional dependence discrimination (ROC-AUC = 0.56). To facilitate practical implementation, we developed a free web-based application that can inform personalized discussions about the potential outcomes of treatment.

Younger age and higher preoperative KPS were associated with better functional outcomes. Studies predicting short-term postoperative functional status also showed age and preoperative KPS among the most relevant predictors.^13,26^ Additionally, the model shows a smaller contrast-enhancing volume association with more favorable functional outcomes. Clinically, the enhancing component may reflect active tumor aggressiveness and represents the target for gross macroscopic resection, as the complete resection of it has been associated with improved survival.^27^ However, a study predicting short-term KPS found overall tumor volume to be only a moderate predictor.^26^ Perhaps the difference in findings stems from disaggregating tumor volume into components, suggesting that contrast-enhancing active tumor, rather than overall tumor size, drives postoperative prognosis. Furthermore, greater cognitive network tumor overlap was associated with functional independence, potentially reflecting the inverse correlation with sensory-limbic involvement. This aligns with previous findings^26^.

The inclusion of MRI-based predictors did not improve prognostic performance beyond the clinical model alone. Although previous studies emphasize the value of multimodal modeling, these typically incorporate intraoperative or postoperative predictors, which may explain the stronger multimodal signal reported.^13,14^ Nevertheless, gains remain modest and short-term functional outcome appears primarily driven by clinical variables, consistent with our findings. The limited added value of MRI-based predictors may reflect their generally limited reproducibility^28^, or instability resulting from inter-observer tumor segmentation variability across tumor sites.^29^ Moreover, different data modalities relevant to this outcome may not be captured by MRI-based characteristics.

Multifocality seems to be the least important prediction of the one-year functional status. Notably, as studies report multifocal gliomas to be more aggressive, have higher proliferation rates, and patients have a significantly shorter overall survival.^30,31^ The contradictory findings here might be attributable to the low prevalence of multifocality in this patient cohort (5%), which results in sparse contributions across samples despite potentially strong effects in a small patient subset. Additionally, multifocality here was extracted by the GSI-RADS toolbox^11^, which defines it as a binary variable based on the disconnected voxel clusters in the automated segmentation counted as separate foci. Unlike the clinical definition^30,31^, this automated approach does not distinguish multifocal or multicentric glioma, omits spread pattern assessment, and remains sensitive to segmentation errors.

Given glioma’s molecular and clinical heterogeneity, making a single confident prediction is inherently misleading. Beyond point predictions, MCP can quantify uncertainty, allowing clinicians to understand how confident the model prediction is. We applied MCP as an exploratory add-on that flagged genuinely unpredictable cases, encouraging clinicians to integrate additional information beyond the three predictors used in the model and to monitor these patients closely over time rather than acting on a falsely confident prediction.

The following limitations should be considered. First, as a single-center study without external validation, generalizability remains limited, though institutional variation in surgical and postoperative protocols would constrain transferability regardless. The heterogeneity in patient characteristics and imaging acquisition across scanners and time periods might add to the model robustness. Second, predictive performance for the minority class of functional dependence was lower, likely reflecting class imbalance inherent to the one-versus-all classifier setup.^24^ Third, postoperative KPS interrater variability may have introduced measurement noise, although the effect is likely minimal given the use of grouped (KPS 10-60 and KPS ≥ 70) rather than exact KPS scores. Finally, restricting inclusion to supratentorial contrast-enhancing tumors excluded glioma mimics such as metastases, abscesses, and lymphomas, while the broad enhancement threshold introduced diagnostic heterogeneity that may improve generalizability across glioma subtypes.

Future studies may extend to other populations, predictors, outcomes, or treatment decisions. Specifically, extending models to biopsy and non-enhancing tumors would enable comparison of functional outcomes across surgical strategies. Moreover, such models might benefit from the inclusion of additional predictors, such as socioeconomic factors^32^, relationship status^33^, or comorbidities^34^. Tumor overlap with cognitive networks favored functional independence, which might indicate that network related predictors could have prognostic value. However, as functional performance is widely distributed across the brain, it might be challenging to establish a clear structure-function relationship.^35^ Furthermore, alternative outcome definitions could include varying KPS thresholds, modeling functional status change over time, or implementing neuropsychological testing for a more nuanced characterization of the onco-functional balance. Finally, exploring associations between long-term functional status and treatment, such as type of treatment, participation in clinical trials, and type of interventions at progression, could provide further insights into factors influencing recovery and inform personalized management strategies.

In conclusion, using a large patient cohort we address the one-year postoperative functional status in contrast-enhancing glioma using only preoperative predictors. We demonstrate that an ML-based prediction of long-term functional status is challenging yet feasible. Integrating such prediction models as supportive tools into the clinical routine likely provides valuable additional information for treatment planning. This study lays the groundwork for future multi-institutional collaboration to externally validate this approach.

## Ethics

Approval was received from the institutional medical ethical committee of the VUmc (Vumc_2021-0437). Informed consent was waived.

## Authorship

Conception and design: E Koderman, L Douw, PC De Witt Hamer, MWA. Caan, PK Tewarie; IMAGO Database management: VC Keil, IJHG Wamelink; Data collection and preparation: MR van Lingen, FB Tijhuis; Model development and analysis scripts: E Koderman; Code review: S Dam; Manuscript preparation: E Koderman; Review of the manuscript: All authors.

## Data Availability

Anonymized data not published within this article will be made available by request from any qualified investigator.

## Acknowledgments

Brigit Thomassen and Cylemarijne Peul contributed to the one-year postoperative KPS assignment. ChatGPT (GPT-5.3 Instant, GPT-4o mini) and Claude (Sonnet 4.5) were used interchangeably for code syntax error resolution and stylistic improvement of language for parts of the introduction and methods. Notebook LM was used to support the review of selected literature. The AI-generated content was reviewed and edited, and the authors take full responsibility for the content of the published article. Some icons on the graphical abstract were obtained from BioRender, but the figure itself was made in PowerPoint.

