## Supplementary material for "Predicting one-year postoperative functional status in contrast-enhancing glioma"

**Tables**

**Supplementary Table 1: Additional patient characteristics.**

| Characteristic | All  (N = 552) | Mortality  (N = 221; 40%) | Functional dependence  (N = 68; 12%) | Functional independence  (N = 263; 48%) |
| --- | --- | --- | --- | --- |
| Sex (females), n (%) | 206 (37) | 86 (39) | 22 (32) | 98 (37) |
| Preoperative seizures,  n (%) | 275 (50) | 91 (41) | 41 (60) | 143 (54) |
| T2 hyperintensity volume (ml), median (Q1-Q3) | 67.9  (33 – 112) | 71.2  (37 – 116) | 60  (31 – 108) | 63.9  (28 – 110) |
| Necrotic core volume (ml), median (Q1-Q3) | 11  (3 – 26) | 13  (5 – 26) | 5  (2 – 22) | 10  (3 – 27) |
| Multifocality, n (%) | 30 (5) | 14 (6) | 7 (10) | 9 (3) |
| Expected resection percentage, n (%)  low / medium / high | 219 (40) /  234 (42) /  99 (18) | 79 (36) /  106 (48) /  36 (16) | 29 (43) /  25 (37) /  14 (21) | 111 (42) /  103 (39) /  49 (19) |
| White matter tracts overlap (%), median (Q1-Q3) | 179  (114 – 215) | 171  (116 – 216) | 172  (109 – 209) | 184  (114 – 215) |
| Sensory limbic overlap (%), median (Q1-Q3) | 15  (9 – 22) | 15  (10 – 21) | 19  (11 – 26) | 14  (8 – 22) |
| Parietal overlap (%), median (Q1-Q3) | 8  (0 – 38) | 11  (2 – 36) | 9  (0.2 – 42) | 6  (0 – 39) |
| Occipital overlap (%), median (Q1-Q3) | 0  (0 – 0.5) | 0  (0 – 0.6) | 0  (0 – 2) | 0  (0 – 0.1) |
| Temporal overlap (%), median (Q1-Q3) | 16  (0.4 – 49) | 21  (1 – 49) | 23  (0.6 – 56) | 12  (0.1 – 48) |
| Frontal overlap (%), median (Q1-Q3) | 19  (7 – 74) | 18  (8 – 66) | 11  (4 – 64) | 21  (5 – 78) |

**Supplementary Table 2: Scan availability across subjects (N total subjects = 552).**

| Scan types availability | T1-weighted with gadolinium contrast | T1-weighted | T2-weighted | Fluid-attenuated inversion recovery | All types of scans available |
| --- | --- | --- | --- | --- | --- |
| Subject count | 552 | 516 | 502 | 513 | 496 |

**Supplementary Table 3:** **Retrospective verification of treatment procedures (N total = 552).**

| Treatment | Count |
| --- | --- |
| Chemotherapy only (%) | 18 (3) |
| Radiotherapy only (%) | 43 (8) |
| Both treatments (%) | 424 (77) |
| No treatment (%) | 68 (12) |
| Passed away before 12m (%) | 61 (90) |
| Astrocytoma or oligodendroglioma (%) | 4 (6) |
| Glioblastoma and alive longer than 12m (%) | 3 (4) |

**Supplementary Table 4: Model performance on the development dataset (N = 386) across the models using different sets of predictors showing mean (± standard deviation) across the 10 folds.** Global (F1 weighted, mean absolute error, quadratic weighted kappa, Matthew’s correlation coefficient) and class specific metrics (ROC-AUC) are shown. Abbreviations: ROC-AUC = receiver operating characteristic – area under the curve.

| Model | F1 weighted | Mean absolute error | Quadratic weighted kappa | Matthew’s correlation coefficient | ROC-AUC  Mortality | | ROC-AUC  Functional dependence | ROC-AUC  Functional independence |
| --- | --- | --- | --- | --- | --- | --- | --- | --- |
| Multimodal | 0.57  ± 0.07 | 0.63  ± 0.11 | 0.4  ± 0.13 | 0.29  ± 0.10 | | 0.73  ± 0.07 | 0.56  ± 0.13 | 0.74  ± 0.08 |
| Clinical | 0.55  ± 0.07 | 0.66  ± 0.11 | 0.36  ± 0.11 | 0.29  ± 0.12 | | 0.75  ± 0.06 | 0.60  ± 0.14 | 0.75  ± 0.07 |
| Top four | 0.58  ± 0.07 | 0.61  ± 0.12 | 0.42  ± 0.14 | 0.31  ± 0.12 | | 0.76  ± 0.06 | 0.60  ± 0.14 | 0.75  ± 0.04 |
| Top three | 0.58  ± 0.06 | 0.61  ± 0.10 | 0.42  ± 0.12 | 0.32  ± 0.09 | | 0.75  ± 0.05 | 0.62  ± 0.15 | 0.76  ± 0.07 |

**Supplementary Table 5:** **Model performance evaluation on the development set using the top three predictors.** Abbreviations: ROC-AUC = Receiver operating characteristic – area under the curve.

| Metric | Development set  (N = 386) |
| --- | --- |
| F1 score |  |
| Mortality | 0.63 |
| Functional dependence | 0.25 |
| Functional independence | 0.63 |
| ROC-AUC |  |
| Mortality | 0.75 |
| Functional dependence | 0.62 |
| Functional independence | 0.76 |
| Quadratic weighted kappa | 0.42 |
| Matthew’s correlation coefficient | 0.32 |

**Supplementary Table 6:** **Mondrian conformal prediction calibration and validation results**. The target coverage for all classes with α = 0.15 was 85%.

| Class | Calibration count  (N = 386) | Calibrated threshold | Minimum probability to be included in the set | Validation count  (N = 166) | Validated coverage |
| --- | --- | --- | --- | --- | --- |
| Mortality | 155 | 0.807 | 19% | 66 | 91% |
| Functional dependence | 48 | 0.852 | 15% | 20 | 85% |
| Functional independence | 183 | 0.797 | 20% | 80 | 80% |

**FIGURES**

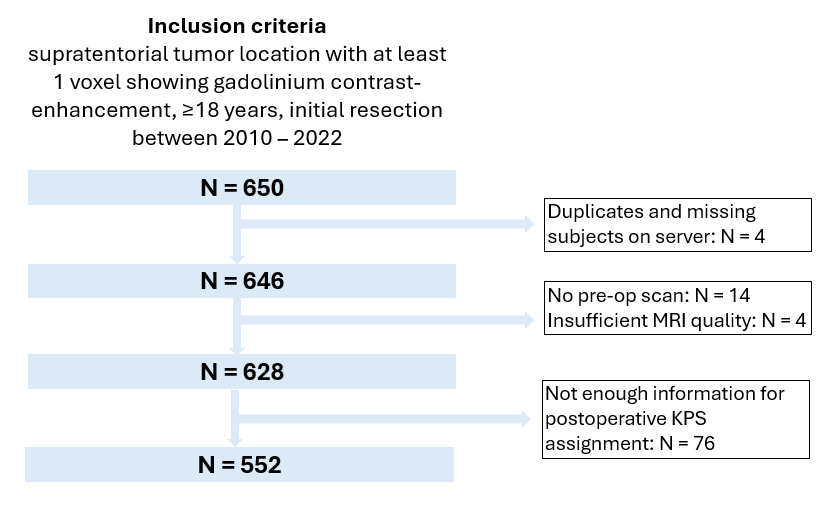

**Supplementary Figure 1:** Data exclusion steps.

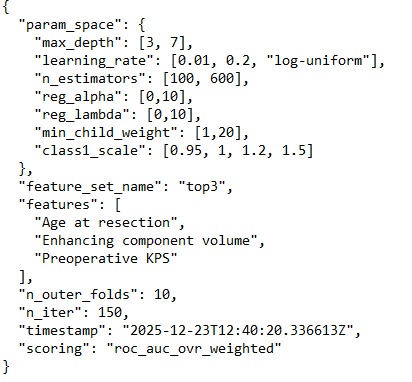

**Supplementary Figure 2:** Hyperparameter tuning details.

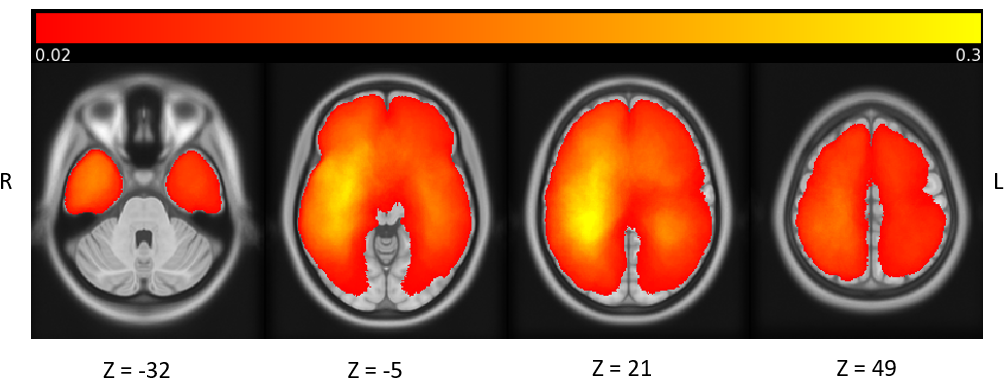

**Supplementary Figure 3:** Spatial tumor mask overlap across all subjects (N = 552). The maps were normalized across the number of patients.

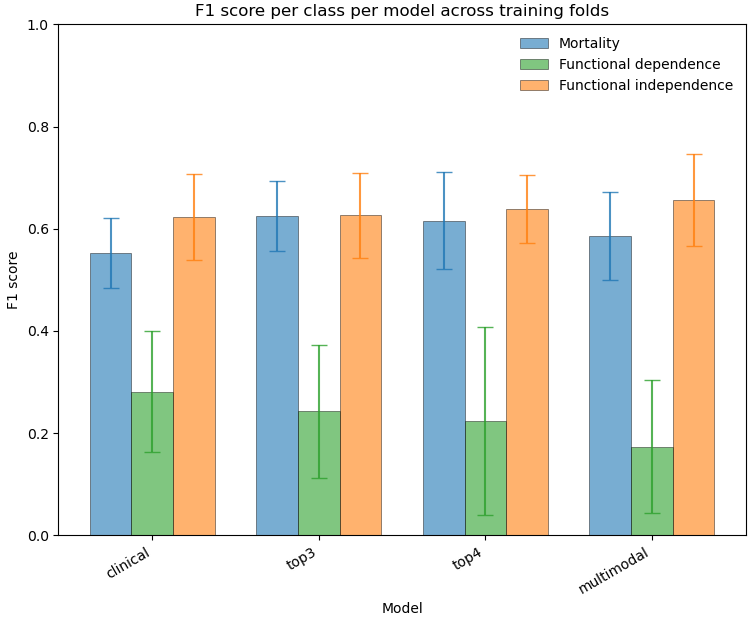

**Supplementary Figure 4:** F1 score per class per model shown in mean with standard deviation across folds.

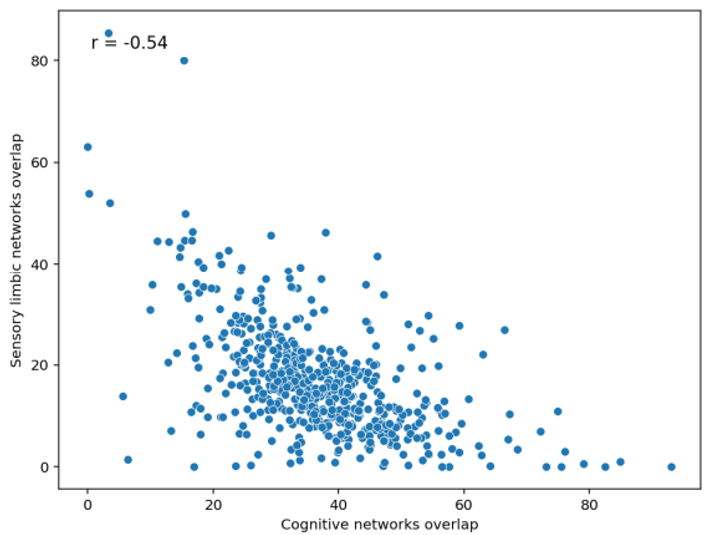

**Supplementary Figure 5:** Negative correlation of tumor overlap (%) with cognitive networks and sensory limbic networks.

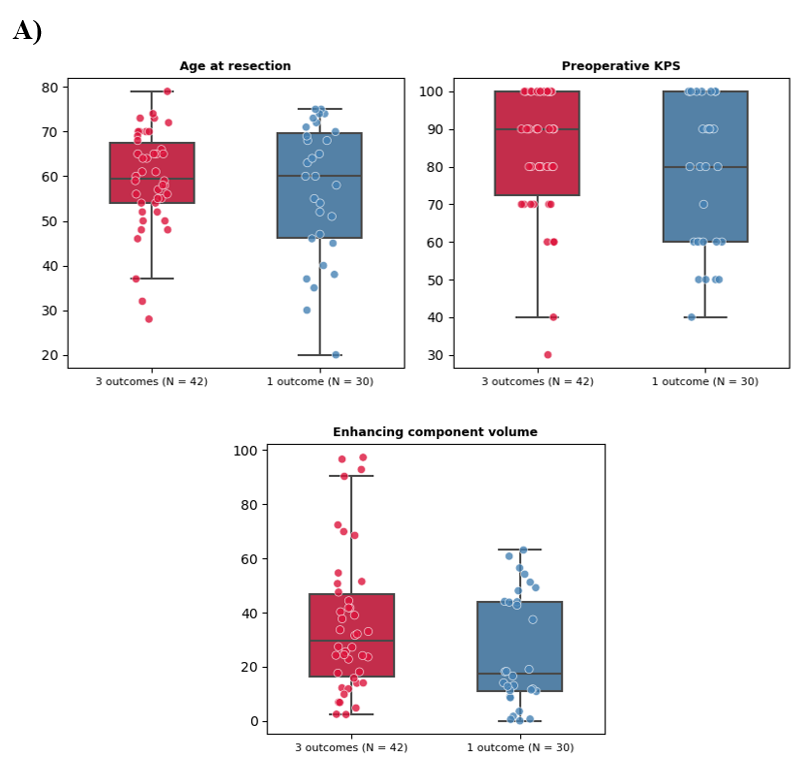

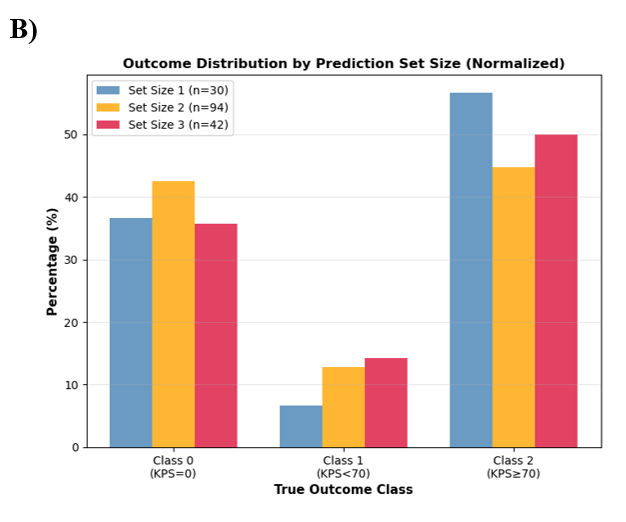

**Supplementary Figure 6:** A) Feature space in the Mondrian conformal prediction comparing patient characteristics in the sets containing three outcomes (unable to successfully discriminate between the 3 outcomes given these predictors) and 1 outcome (clear prognosis). B) Outcome distribution by prediction set size normalized by the count of each outcome within a given set size (certain, moderate certain, uncertain) divided by total number of predictions in this group. Uncertainty is evenly distributed and not biased towards a certain outcome. Balanced outcomes reflect equiprobable cases.
